# The Role of Deep Learning in Diagnostic Imaging of Spondyloarthropathies: A Systematic Review

**DOI:** 10.1101/2024.05.15.24307396

**Authors:** Mahmud Omar, Abdulla Watad, Dennis McGonagle, Shelly Soffer, Benjamin S Glicksberg, Girish N Nadkarni, Eyal Klang

**Affiliations:** Tel-Aviv University, Faculty of Medicine. Tel-Aviv, Israel; Department of Medicine B and Zabludowicz Center for Autoimmune Diseases, Sheba Medical Center, Tel-Hashomer, Ramat-Gan 5265601, Israel; Section of Musculoskeletal Disease, NIHR Leeds Musculoskeletal Biomedical Research Centre, Leeds Institute of Rheumatic and Musculoskeletal Medicine, University of Leeds, Chapel Allerton Hospital, Leeds, UK; Institute of Hematology, Davidoff Cancer Center, Rabin Medical Center, Petah-Tikva, Israel; The Charles Bronfman Institute of Personalized Medicine, Icahn School of Medicine at Mount Sinai, New York, New York, USA; Division of Data-Driven and Digital Medicine (D3M), Icahn School of Medicine at Mount Sinai, New York, New York, USA

## Abstract

**Aim:** Diagnostic imaging is an integral part of identifying spondyloarthropathies (SpA), yet the interpretation of these images can be challenging. This review evaluated the use of deep learning models to enhance the diagnostic accuracy of SpA imaging.

**Methods:** Following PRISMA guidelines, we systematically searched major databases up to February 2024, focusing on studies that applied deep learning to SpA imaging. Performance metrics, model types, and diagnostic tasks were extracted and analyzed. Study quality was assessed using QUADAS-2.

**Results:** We included 22 studies demonstrating that deep learning aids in diagnosing and classifying SpA, differentiating arthritis forms, and estimating disease progression and structural changes. These models, particularly those using advanced U-Net architectures, consistently outperformed traditional diagnostic methods, showing a notable increase in diagnostic accuracy.

**Conclusion:** Deep learning models are excellent for augmenting the accuracy of SpA imaging diagnostics. Despite their potential, challenges in overcoming retrospective study biases and integrating these models into clinical practice remain. Future directions should aim to validate these models in real-world clinical settings.

## Introduction

Medical imaging, including X-rays and Magnetic Resonance Imaging (MRI), plays a vital role in diagnosing Spondyloarthropathies (SpA) (1–3). Despite the importance of imaging, interpretating these images is challenging due to the subtle and variable manifestations of SpA (1).

In recent years, artificial intelligence (AI), particularly deep learning, has shown promise in enhancing diagnostic accuracy across various medical fields, including rheumatology and radiology (2–5) . Deep learning employs artificial neural networks designed to analyze images efficiently. Its application in diagnosing SpA, however, has not been comprehensively reviewed (2,6). Convolutional Neural Networks (CNNs), a type of deep learning method, have notably improved diagnostic accuracy in medical imaging, including for SpA (7–11). Unlike classical machine learning, which relies on manual feature extraction, deep learning automates this process, enabling more accurate interpretations of radiological images (12) (**Figure 1**).

**Figure 1.**
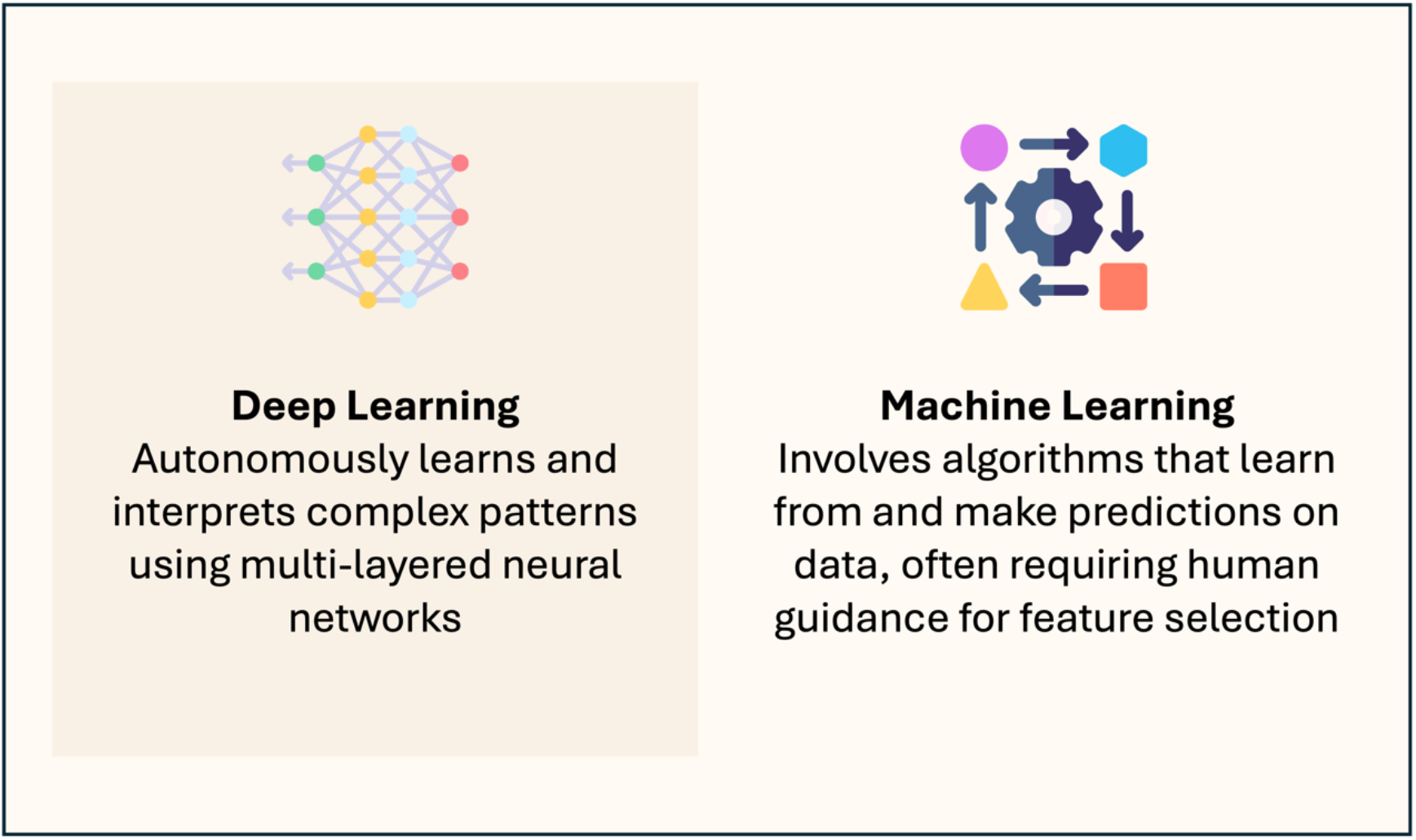
Comparative Overview of Deep Learning and Machine Learning.

In this systematic literature review, we assess the effectiveness of deep learning techniques in enhancing the accuracy of diagnostic imaging for SpA. Our goal is to determine how these advanced models contribute to the precision of imaging interpretations in the clinical setting.

### Fundamentals Concepts of Deep Learning and Computer Vision

#### Computer Vision Tasks in Medical Imaging

Three essential computer vision tasks—classification, detection, and segmentation— use deep learning algorithms to improve diagnostic precision (13,14).

*Classification* assigns each image to a specific category based on established criteria. In the context of SpA, deep learning models can distinguish between normal, inflammatory, and structural changes. This task is fundamental for assessing disease stage, guiding treatment decisions, and evaluating treatment effectiveness (13,14).

*Detection* involves locating key features in medical images and marking them with a region of interest (ROI) (10). For SpA, this task may target the identification of inflammation or structural changes such as erosions or fusions in the spine and sacroiliac joints. Deep learning algorithms scan images and visually mark significant abnormalities, aiding clinicians in quickly identifying critical areas (13,14).

*Segmentation* partitions a digital image into segments by delineating the exact pixel-wise borders of areas of interest, such as lesions or organs. In SpA, segmentation may accurately outline affected areas in the joints or spine, thereby enabling measurement of the extent of inflammation or bone growth. This precision is essential for assessing disease severity and monitoring progression (10,13,14).

#### Deep Learning Models in Medical Imaging

Currently, in the computer vision field, the primarily used deep learning algorithm is convolutional neural networks (CNN) (5,15). This methodology excels in image analysis by identifying repeating patterns through a multi-layered approach (15) (**Figure 2**). U-Net, a subset of CNN, specializes in segmenting medical images to precisely highlight areas of interest (16), such as borders of an inflammatory process (17). CNNs include several types of models such as ResNet, EfficientNet and others. These models can analyze intricate details in images, including complex 3-dimensional, multi-anatomical planes (**Figure 3**) (18–21).

**Figure 2:**
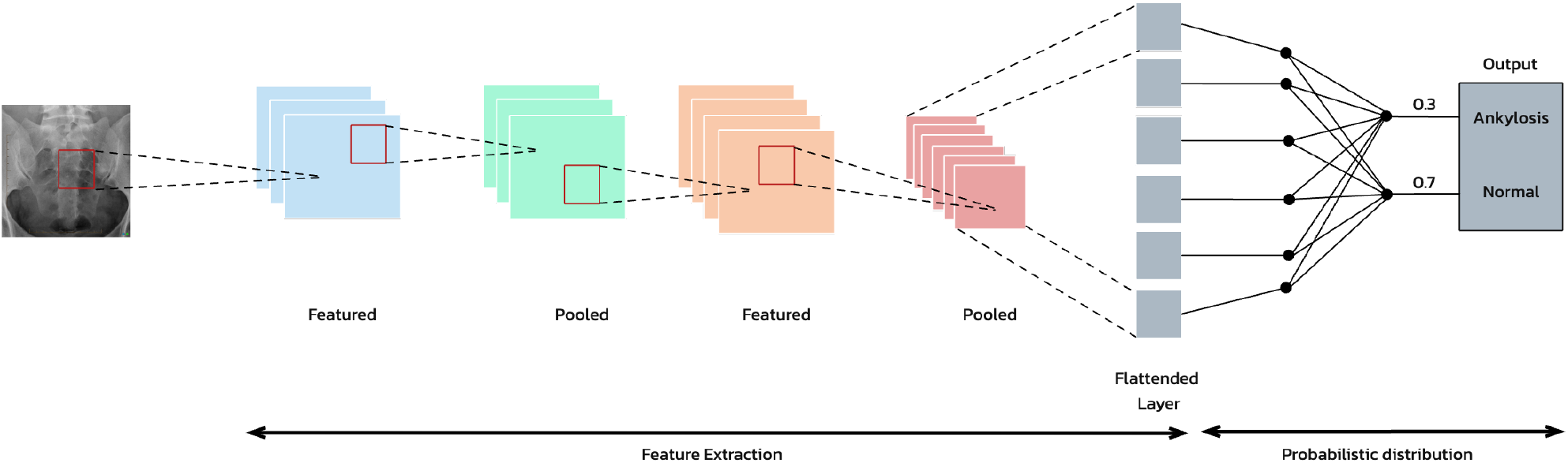
Typical Convolutional Neural Networks (CNN) Workflow for the diagnosis of ankylosis. This schematic represents a convolutional neural network (CNN) architecture used for medical image analysis. An MRI input undergoes preprocessing to enhance features, which is then passed through consecutive convolutional layers where features are extracted and analyzed. Each layer applies filters to detect specific attributes, pooling layers to reduce dimensionality, and activation functions to introduce non-linearity, facilitating the identification of complex patterns. The final classification is done through fully connected layers, resulting in the output that categorizes the image into diagnostic categories, such as ‘Ankylosis’ or ‘Normal’.

**Figure 3:**
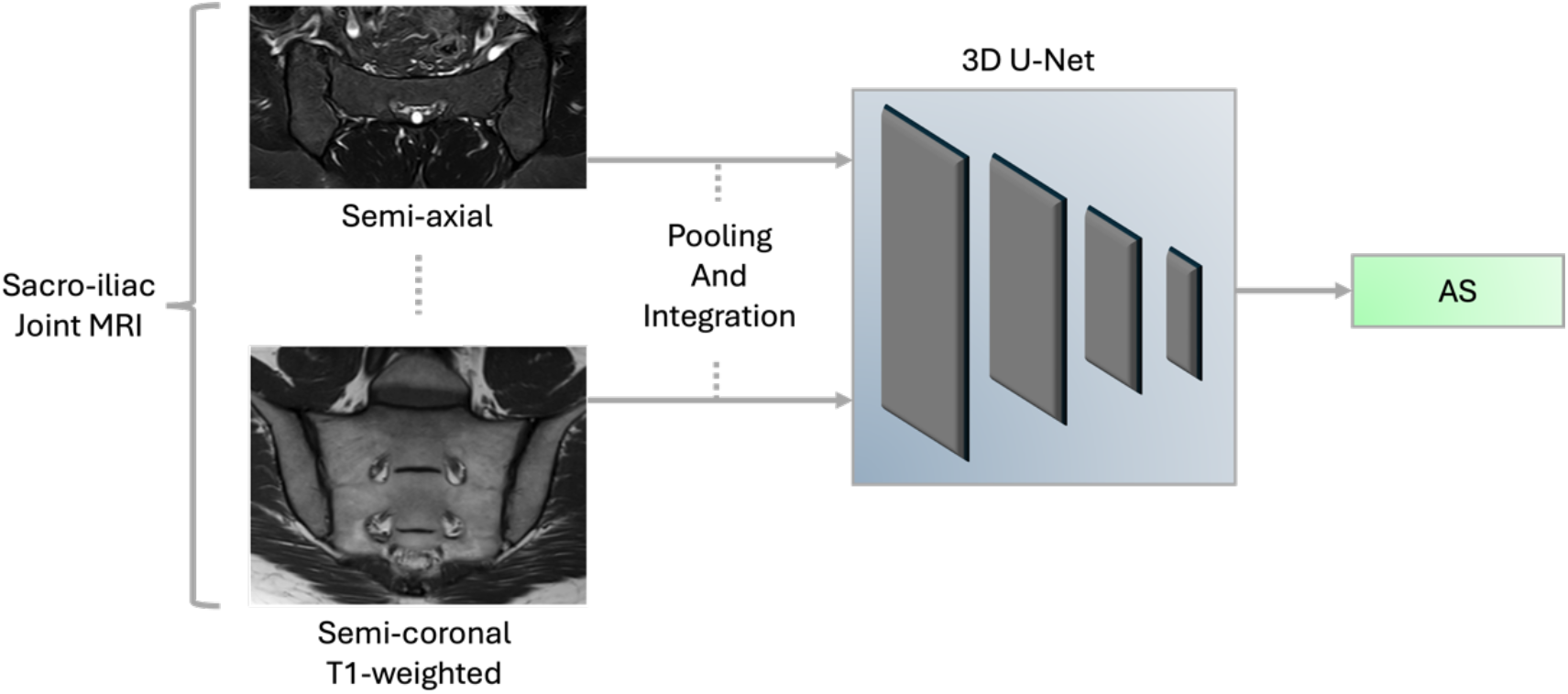
Deep Learning Analysis of MRI for AS Diagnosis Utilizing 3D-Unet. This figure presents a deep learning framework, specifically a U-Net model, processing MRI scans to diagnose Ankylosing Spondylitis (AS). Two MRI inputs, representing different anatomical planes, feed into the neural network which then utilizes convolutional layers to extract features and identify patterns indicative of AS, leading to a diagnostic output.

These models have transformed radiology, enhancing diagnostic accuracy and efficiency in interpreting a wide range of imaging modalities (2,12,16,18,22,23). These techniques have also proven effective in detecting subtle signs of SpA, differentiating it from other conditions, and aiding in the assessment of disease progression (7,8,21,24).

## Methods

### Search Strategy

The review was registered with the International Prospective Register of Systematic Reviews - PROSPERO (Registration code: CRD42024517372). We adhered to the Preferred Reporting Items for Systematic Reviews and Meta-Analyses (PRISMA) guidelines (25,26). A systematic search of key databases including PubMed, Embase, Web of Science, and Scopus was conducted, concluding in February 2024. To enhance our search, we supplemented our database inquiries with manual screening of the references of included studies and searches on Google Scholar. Our search strategy employed a combination of specific keywords related to artificial intelligence such as ‘Artificial Intelligence,’ ‘Deep Learning,’ ‘Neural Networks’, along with terms pertinent to spondyloarthritis, and terms relating to diagnostic imaging techniques including ‘MRI,’ ‘CT,’ and ‘X-ray.’ Detailed search strings for each database are provided in the **Supplementary Materials**.

### Study Selection

We included original research articles that focused on the integration of deep learning in the radiographic diagnosis of SpA. Studies were selected if they provided data for assessing the performance metrics of models, such as area under the curve, accuracy, sensitivity, and specificity. We excluded review papers, case reports, conference abstracts, editorials, preprints, and studies not conducted in English.

### Data Extraction

Two independent reviewers [initials] extracted data from each study using a structured form, ensuring coverage of relevant variables. These included the title, author, publication year, study design, radiology method used, the body part examined, research task, sample size, AI method/model employed, performance metrics, limitations, main results, and their implications. Any differences in data extraction were resolved through collaborative discussion, with a third reviewer’s input sought when necessary.

### Risk of Bias

To evaluate the quality and robustness of the methodologies in the included studies, the QUADAS-2 (Quality Assessment of Diagnostic Accuracy Studies-2) tool was used (27).

## Results

### Search Results and Study Selection

Our search across PubMed, Embase, Web of Science, and Scopus initially identified 897 papers. After removing 472 duplicates, 425 articles remained. Subsequent title and abstract screening excluded 354 papers, leaving 71 full-text articles for evaluation. Ultimately 21 articles were selected for inclusion. One additional article was identified through reference checking, leading to a total of 22 studies included in our review (6–9,17,19–21,24,28–40). The selection process is visually represented in **Figure 4**, the PRISMA flowchart.

**Figure 4.**
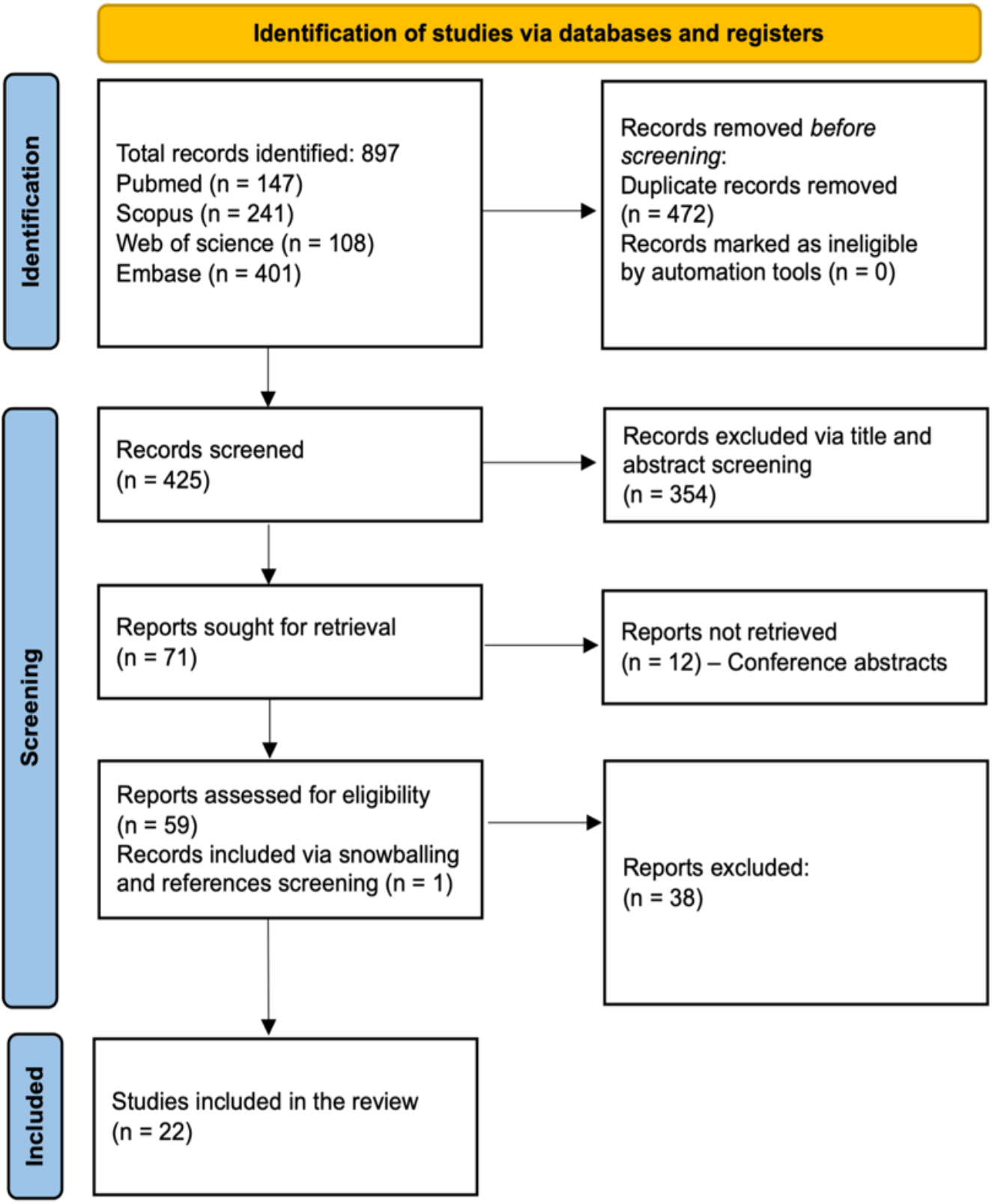
PRISMA flowchart.

### Risk of Bias

The risk of bias analysis (**Figure S1)** highlighted that most studies demonstrated a low risk across various domains. Specifically, seven studies exhibited low risk in all evaluated domains, reinforcing their credibility and methodological robustness (6,8,21,28,31,36,40). Overall, 17 studies were classified as low risk (6–8,19– 21,24,28–34,36,37,40), three as high risk (17,35,38), and two as presenting some concerns (9,39) (**Figure S1-2**), underscoring a generally low risk of bias. Regarding applicability concerns, most studies showcased a low risk, indicating their relevance to broader clinical settings. However, it is noteworthy that some studies faced limitations due to small sample sizes.

### Overview of Included Studies

This systematic review includes 22 studies exploring the use of deep learning in SpA imaging diagnosis, published between 2019 and 2024(6–9,15,17–20,26–38) (Figure S3). These studies, predominantly published in impactful quartile 1 (Q1) journals **(Figure 5, Table S1)**.

**Figure 5.**
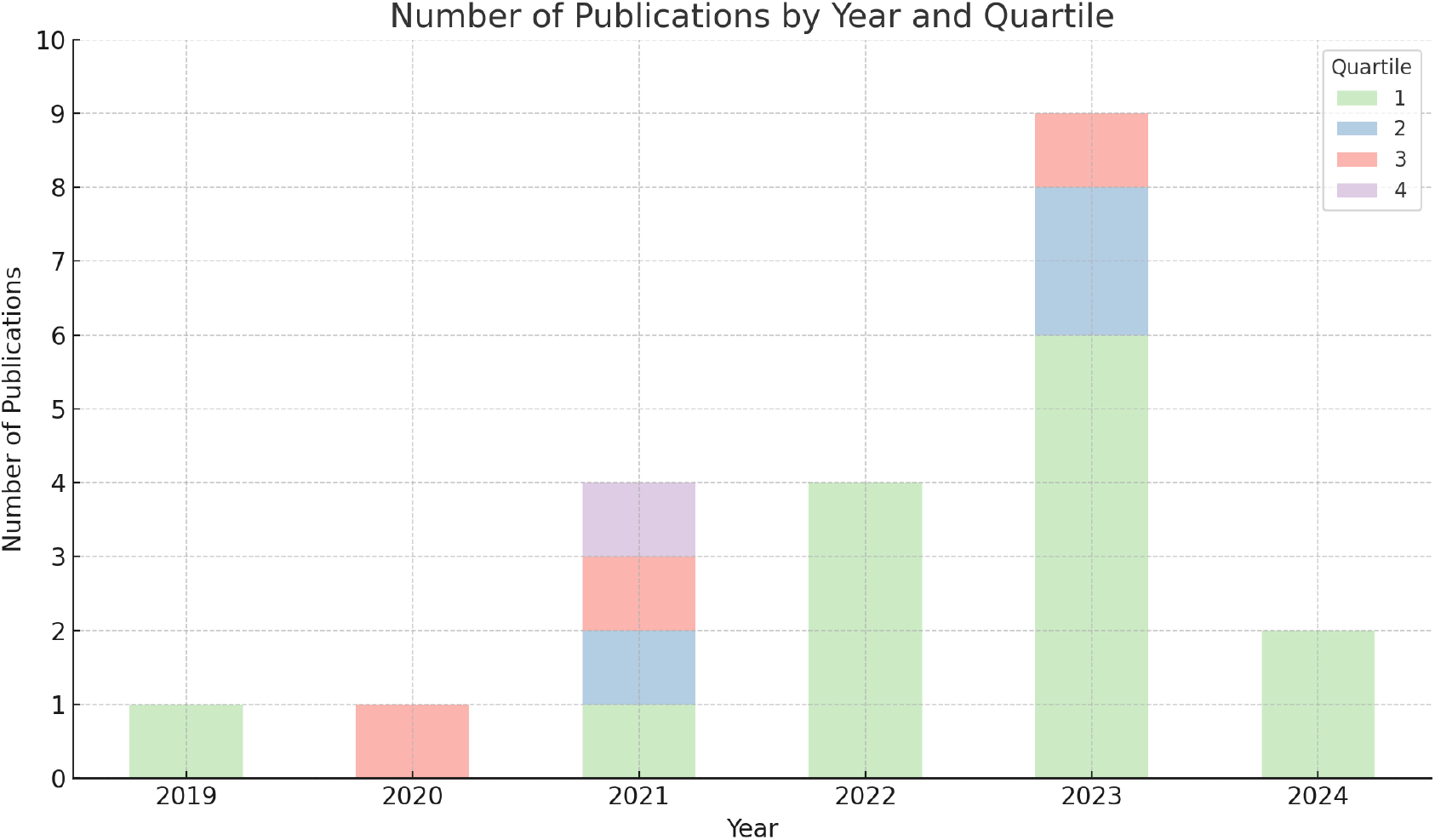
Distribution of the Included Studies by Quartile and Year.

The studies involved diverse patient populations. Sample sizes range from smaller cohorts, such as 56 patients in Faleiros et al. (28), to more extensive datasets, like the 6436 pelvic X-rays analyzed in Li et al. (7).The clinical tasks implemented were also diverse, addressing different aspects of SpA diagnosis and management. Examples include the detection of inflammatory sacroiliitis (19,38), differentiating arthritis types (20), grading vertebral changes (39), and predicting the course of ankylosing spondylitis (7).

The radiographic images analyzed in this review included X-ray, MRI, and Computed Tomography (CT). Specifically, MRI was employed in 12 studies, with a focus on the sacroiliac joint and the whole spine. Five studies utilized CT, while another five employed X-ray, primarily for diagnosing sacroiliitis and grading vertebral changes. A wide array of models has been employed. For instance, Lee et al. (2023) used a two-stage framework combining Faster R-CNN and VGG-19 (19), while Bressem et al. employed a sophisticated 3D U-Net architecture followed by a dual-encoder ResNet-101 (40). Koo et al. utilized a modified HRNet for key-point detection alongside a ResNet 152-based CNN (39).

The performance of these models was generally promising, though varied. Sensitivity, specificity, and AUROC (Area Under the Receiver Operating Characteristic Curve) are commonly used metrics. For example, Lee et al. reported a patient-level sensitivity of 94.7% and specificity of 69.1%, with an AUC of 0.816 (19). Bressem et al. achieved an AUC of 0.94 for detecting inflammatory changes (40). Koo et al. demonstrated high sensitivity 93.6% and accuracy 95.7% for grading vertebral changes in ankylosing spondylitis (39). (**Figure S3** summarizes the most important performance metrics in the studies that reported sensitivity, specificity, or AUC).

### Clinical tasks

The clinical tasks of the included studies can be grouped into three broad categories: *Diagnosis and Classification of SpA, Differentiation of Arthritis Forms*, and *Analysis of Disease Progression and Structural Changes* (**Figure S4** visually illustrates the distribution of the clinical tasks among the included studies). Each category encapsulates the key findings, performance metrics, and notable characteristics of the AI models utilized in the studies.

### Diagnosis and Classification of SpA

*In MRI studies*, Lee et al. (2023 - MRI), Bressem et al. (2022 - MRI), Bordner et al, and Roles et al. focused on diagnosing axial SpA (19,37,38,40).

Lee et al. utilized a two-stage faster R-CNN and VGG-19 network achieving a sensitivity of 94.7% and specificity of 69.1% (19). Bressem et al. (2022) reported an AUC of 0.94 with their 3D dual-encoder ResNet-101 for detecting inflammatory changes (40). Bordner et al. developed a mask-RCNN modelpredicting active sacroiliitis with high accuracy, evidenced by MCC values up to 0.90 and AUCs up to 0.98 (36). Roles et al. study utilized a ResNet18-based CNN, achieving a high cross-validation AUC of 94.5% (37). Zhang (2024) et al. and Li et al. demonstrated the potential of CNNs in diagnosing sacroiliitis and ankylosing spondylitis (9,20), with Li’s ensemble models surpassing human experts (precision, recall, and AUC values of 0.91, 0.90, and 0.96, respectively) (7). Faleiros et al. and Lin et al. also showed high accuracy in MRI classification for active inflammatory sacroiliitis and spinal inflammation, respectively, matching radiologist performance (28,29).

*“In X-ray studies*, Lee et al. (2023 – Xray) and Bressem et al. (2021 - Xray) utilized deep learning models to diagnose sacroiliitis from radiographs (8,33). Lee et al. utilized a DenseNet121 CNN, achieving up to 100% sensitivity and specificity for certain grades of spondylolisthesis (33). Bressem et al. (2021), employedResNet-50, with AUC of 0.97 and 0.94 invalidation and test datasets, respectively (8). Additionally, Ureten et al. applied pre-trained models like VGG-16, ResNet-101, and Inception-v3, reaching an accuracy of 89.9% with VGG-16 (21).

*For CT images*, Zhang et al. (2023) utilized nnU-Net and a 3D CNN to grade sacroiliitis in ankylosing spondylitis (17). The nnU-Net achieved Dice coefficient of 0.889 for the test set, indicating high segmentation accuracy. For grading sacroiliitis, the 3D CNN model yielded micro average AUCs of 0.91 for the test set, with accuracy levels of 0.802.

### Differentiation of Arthritis Forms

Folle et al. study on differentiating Psoriatic Arthritis (PsA) from other forms using ResNet neural networks showed AUC values ranging from 67% to 75% (20).

### Disease Progression and Structural Changes Analysis

Five studies examined radiographic progression and structural changes. Koo et al. study utilized a CNN model for grading vertebral bodies changes, showing high sensitivity and accuracy (39). Gou et al. used LHR-Net for lesion segmentation and ankylosing spondylitis grading, with a dice coefficient score of 0.71 (36).

Zhang (2023) et al and Berghe et al. leveraged CT imaging for sacroiliitis analysis (17,34). Zhang et al. nnU-Net achieved a Dice coefficient of 0.915, indicating high segmentation accuracy (17). Berghe et al. study utilized U-Net for sacroiliac joint segmentation with a dice coefficient of 0.75 (34). Lee et al. (2021) utilized ResNet18 to detect bone marrow edema in the sacroiliac joints from MRI images, achieving an impressive accuracy of 93.5% (35).

## Discussion

We analyzed 22 studies that employed deep learning models to enhance the diagnostic imaging of SpA using MRI, CT, and X-ray techniques. Our findings indicate MRI as the most effective modality, highlighted by Lee et al.’s two-stage framework with high sensitivity and specificity (19), and Bressem et al.’s 3D CNN architecture, reaching an AUC of 0.94 (40). These models often surpassed the performance of human experts in diagnosing and classifying axial SpA. CT imaging also demonstrated strong results, especially in segmentation and grading of sacroiliitis, with Zhang et al.’s nnU-Net showing high segmentation accuracy and diagnostic reliability (17). In contrast, X-ray-based models, while still effective, generally showed lower performance compared to MRI and CT.

The integration of CNNs and U-Net has markedly improved the accuracy of imaging diagnoses in SpA, presenting a potential in transformation rheumatology and radiology (6,8,28,29,31). Deep learning methods have excelled in identifying complex patterns and segmenting medical images (2,15,18). However, the performance metrics across studies, such as sensitivity, specificity, and AUC, exhibit variability. This variability underscores the necessity for standardized benchmarks in future research to ensure the consistency and reliability of AI applications.

Integrating AI with human expertise is crucial for nuanced interpretation and decision-making (2,15). For example, the study by Koo et al. demonstrated high sensitivity and accuracy in grading vertebral changes in ankylosing spondylitis using a modified HRNet alongside a ResNet 152-based CNN (39). This integration highlights AI’s potential to augment human diagnostic capabilities, emphasizing the importance of collaborative efforts between technology and clinical expertise (2,3,23,41).

While CNNs, particularly ResNets and U-Nets, were predominantly used, notable exceptions include Tenorio et al.’s use of a fully connected artificial neural network (ANN) for radiomic model development, which yielded high results (31). Additionally, an Attention U-Net algorithm provided a unique adaptation by enhancing the standard U-Net with attention mechanisms for more effective focus on pertinent image areas (22). With the recent advent of attention and transformer models in computer vision, their utilization for SpA is expected to increase in the coming years.

The results of our study, along with the current direction of the literature, underscore that the potential impact of AI’s integration into clinical practice, particularly within the fields of rheumatology and radiology (2–4,42,43). However, prospective studies are crucial to validate AI models, especially those based on deep learning, and to investigate their applicability in the clinical setting (44). The use of AI in diagnostic imaging faces challenges. These include the dependency on data quality and variability in study methodologies (2,23). Ensuring that models accurately reflect the heterogeneity of SpA is critical. Addressing these limitations is pivotal for the safe and effective deployment of AI.

The limitations of the reviewed studies stem from their retrospective nature, often leading to challenges in data diversity and applicability in clinical settings. Additionally, most studies did not compare AI model performance with human practitioners. For instance, Lee et al. relied on expert consensus for ground truth, possibly limiting real-world applicability (19). Folle et al. faced challenges with insufficient training datasets, a common issue in deep learning studies that affects model robustness (18). Similarly, Bressem et al. encountered biases due to their selective use of imaging techniques, potentially affecting the comprehensiveness of their findings (38). These examples highlight common issues in this specific area of AI research, such as data quality, model validation, and potential biases, which are important for the practical application of AI in medical imaging **(Table 2)**. Additionally, a meta-analysis was not conducted due to the heterogeneity of the studies involved (43).

In conclusion, deep learning models offer improvements in the accuracy of SpA imaging diagnostics, particularly with MRI. Despite their potential, transitioning them into clinical application requires overcoming current retrospective biases and integrating AI with clinical expertise. Future directions should aim to standardize methodologies and validate these models in diverse clinical settings.

## Data Availability

All data produced in the present work are contained in the manuscript

## Acknowledgment

None

## Financial disclosure

None

Specific Boolean strings used to screen the different databases:

**PubMed** (“Artificial Intelligence” OR “AI” OR “Deep Learning” OR “Machine Learning” OR “Neural Networks” OR “Computer Vision” OR “Automated Diagnosis” OR “Algorithm*”) AND (“Spondyloarthritis” OR “Ankylosing Spondylitis” OR “Axial Spondyloarthritis” OR “Spinal Arthritis” OR “Spondyloarthropathy” OR “Inflammatory Back Pain”) AND (“Radiography” OR “Radiographic Diagnosis” OR “MRI” OR “Magnetic Resonance Imaging” OR “CT” OR “Computed Tomography” OR “X-Ray” OR “X-Ray Imaging” OR “Radiograph” OR “Imaging Techniques” OR “Diagnostic Imaging”)

**Embase** (‘artificial intelligence’/exp OR ‘AI’ OR ‘deep learning’ OR ‘machine learning’ OR ‘neural networks’ OR ‘computer vision’ OR ‘automated diagnosis’ OR ‘algorithm*’) AND (‘spondyloarthritis’/exp OR ‘ankylosing spondylitis’/exp OR ‘axial spondyloarthritis’ OR ‘spinal arthritis’ OR ‘spondyloarthropathy’ OR ‘inflammatory back pain’) AND (‘radiography’/exp OR ‘radiographic diagnosis’ OR ‘MRI’ OR ‘magnetic resonance imaging’/exp OR ‘CT’ OR ‘computed tomography’/exp OR ‘x-ray’/exp OR ‘x-ray imaging’ OR ‘radiograph’ OR ‘imaging techniques’ OR ‘diagnostic imaging’)

**Web of Science** (TS=(“Artificial Intelligence” OR “AI” OR “Deep Learning” OR “Machine Learning” OR “Neural Networks” OR “Computer Vision” OR “Automated Diagnosis” OR “Algorithm*”)) AND (TS=(“Spondyloarthritis” OR “Ankylosing Spondylitis” OR “Axial Spondyloarthritis” OR “Spinal Arthritis” OR “Spondyloarthropathy” OR “Inflammatory Back Pain”)) AND (TS=(“Radiography” OR “Radiographic Diagnosis” OR “MRI” OR “Magnetic Resonance Imaging” OR “CT” OR “Computed Tomography” OR “X-Ray” OR “X-Ray Imaging” OR “Radiograph” OR “Imaging Techniques” OR “Diagnostic Imaging”))

**Scopus** ( TITLE-ABS-KEY ( “Artificial Intelligence” OR “AI” OR “Deep Learning” OR “Machine Learning” OR “Neural Networks” OR “Computer Vision” OR “Automated Diagnosis” OR “Algorithm*” ) ) AND ( TITLE-ABS-KEY ( “Spondyloarthritis” OR “Ankylosing Spondylitis” OR “Axial Spondyloarthritis” OR “Spinal Arthritis” OR “Spondyloarthropathy” OR “Inflammatory Back Pain” ) ) AND ( TITLE-ABS-KEY ( “Radiography” OR “Radiographic Diagnosis” OR “MRI” OR “Magnetic Resonance Imaging” OR “CT” OR “Computed Tomography” OR “X-Ray” OR “X-Ray Imaging” OR “Radiograph” OR “Imaging Techniques” OR “Diagnostic Imaging” ) )

## Notes

### Competing Interest Statement

The authors have declared no competing interest.

### Funding Statement

This study did not receive any funding

