## Supplementary materials for "The Role of Deep Learning in Diagnostic Imaging of Spondyloarthropathies: A Systematic Review"

**Table S1: Impact Factor and Quartile Classifications of Journals Publishing Included Studies.**

| Quartile | Impact Factor (IF) | Journal | Year | Author |
| --- | --- | --- | --- | --- |
| 1 | 7.6 | Frontiers in Immunology | 2023 | Lee |
| 1 | 5.5 | Rheumatology (Oxford) | 2022 | Folle |
| 1 | 29.1 | Radiology | 2022 | Bressem |
| 1 | 3.65 | Therapeutic advances in musculoskeletal disease | 2022 | Koo |
| 1 | 7.42 | Diagnostic and Interventional Imaging | 2023 | Bordner |
| 1 | 15.4 | Arthritis & rheumatology | 2023 | Roles |
| 4 | 4.1 | Physics in medicine and biology | 2021 | Gou |
| 3 | 3.99 | Diagnostics | 2021 | Lee |
| 1 | 4.5 | European journal of radiology | 2024 | Zhang |
| 1 | 4.9 | Journal of Digital Imaging | 2023 | Zhang |
| 3 | 3.99 | Diagnostics | 2023 | Lee |
| 1 | 7 | European radiology | 2023 | Berghe |
| 1 | 4.9 | Journal of Digital Imaging | 2022 | Tenorio |
| 1 | 10.9 | Medical image analysis | 2019 | Shenkman |
| 1 | 5.6 | Arthritis Research & Therapy | 2021 | Bressem |
| 2 | 3.9 | Frontiers in public health | 2023 | Li |
| 1 | 3 | Biomedicines | 2023 | Tas |
| 2 | 2.86 | Modern rheumatology | 2023 | Ureten |
| 2 | 8.6 | Biocybernetics and Biomedical Engineering | 2021 | Rzecki |
| NA | NA | Lecture Notes on computer science (Book Chapter and Conference article). | 2022 | Fernandez |
| 1 | 2.8 | European Spine Journal | 2024 | Lin |
| 3 | 3 | Advances in Rheumatology | 2020 | Faleiros |

*Impact factors were extracted from the websites of each corresponding journal. Quartiles were extracted from Scimago.

**Risk of bias figures**

**Figure S1: Distribution of Risk of Bias Concerns Across Individual Domains.**

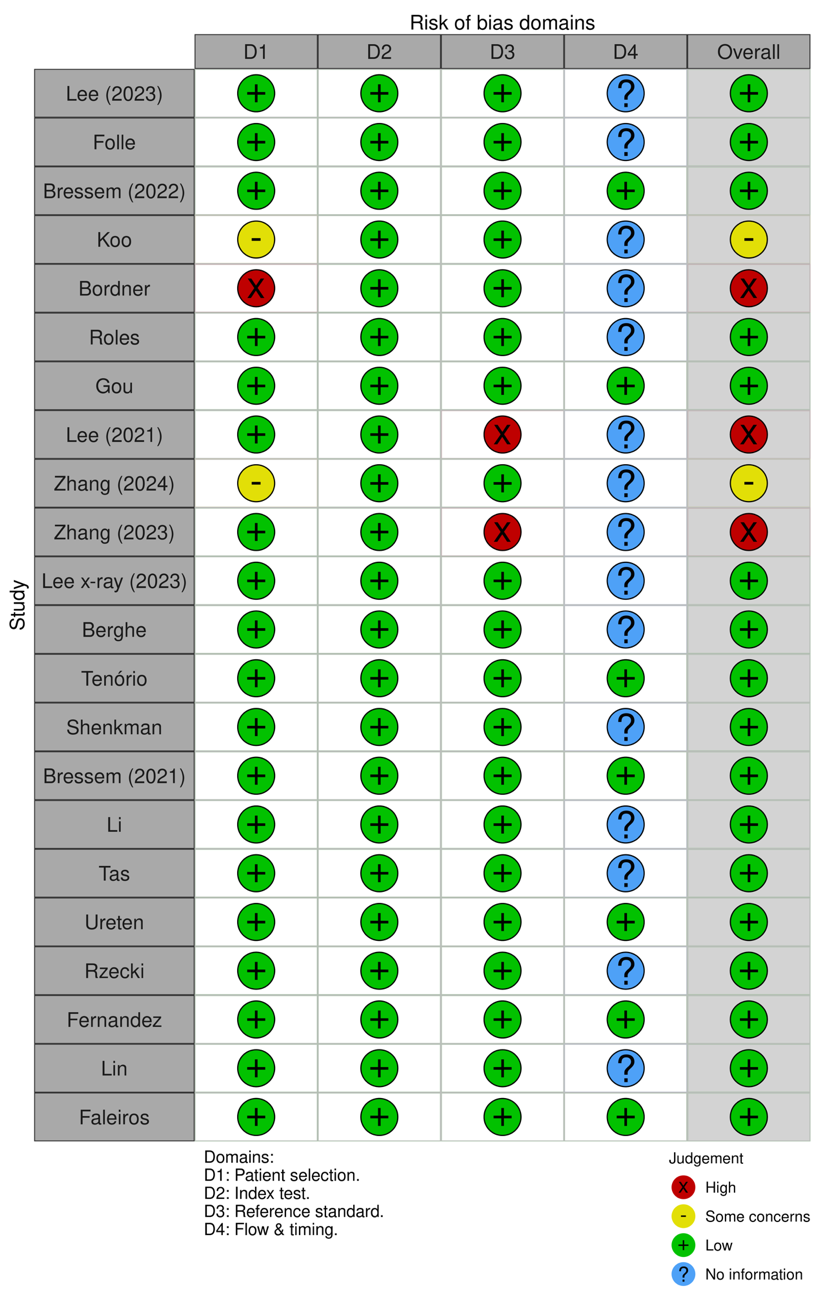

**Figure S2: Cumulative Assessment of Overall Risk of Bias Concerns Across All Included Studies.**

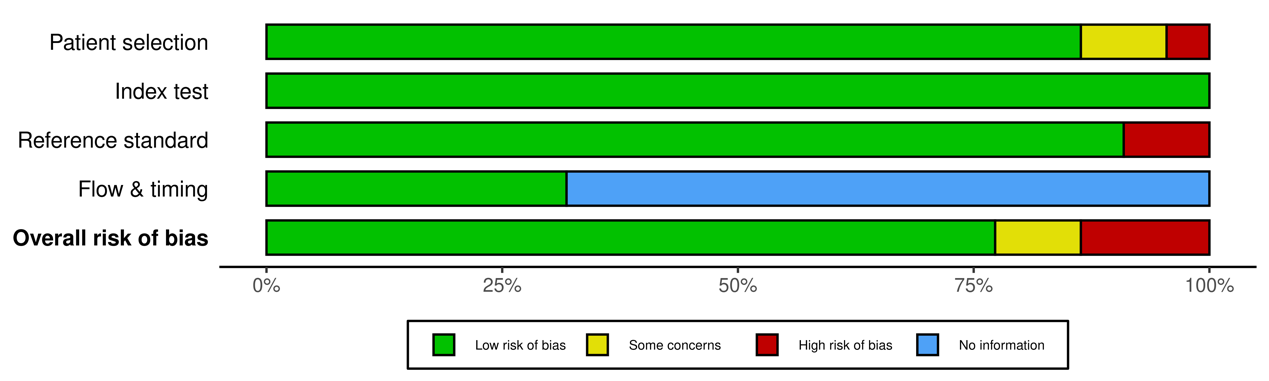

**Additional figures.**

**Figure S3: Bar chart visualizing the deep learning Model Performance Metrics across various studies.**

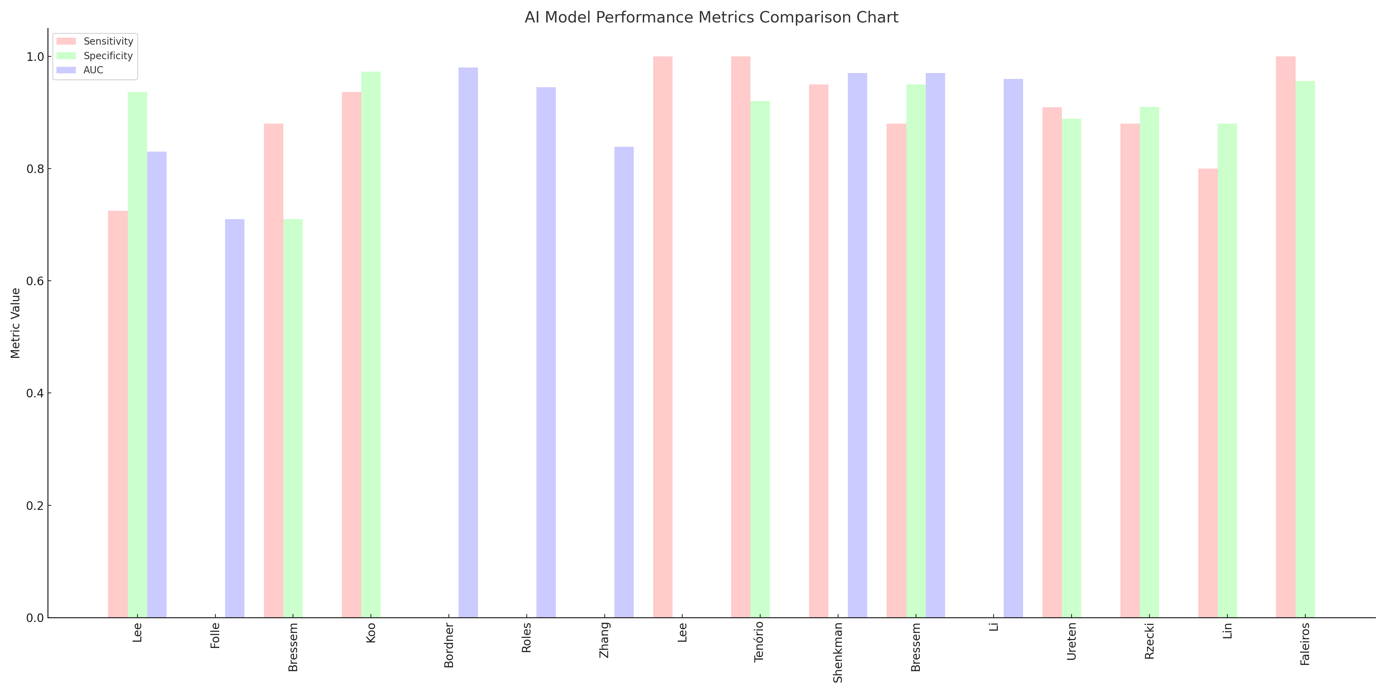

**Figure S4: Distribution of the Clinical Tasks Performed by the Deep Learning Models.**

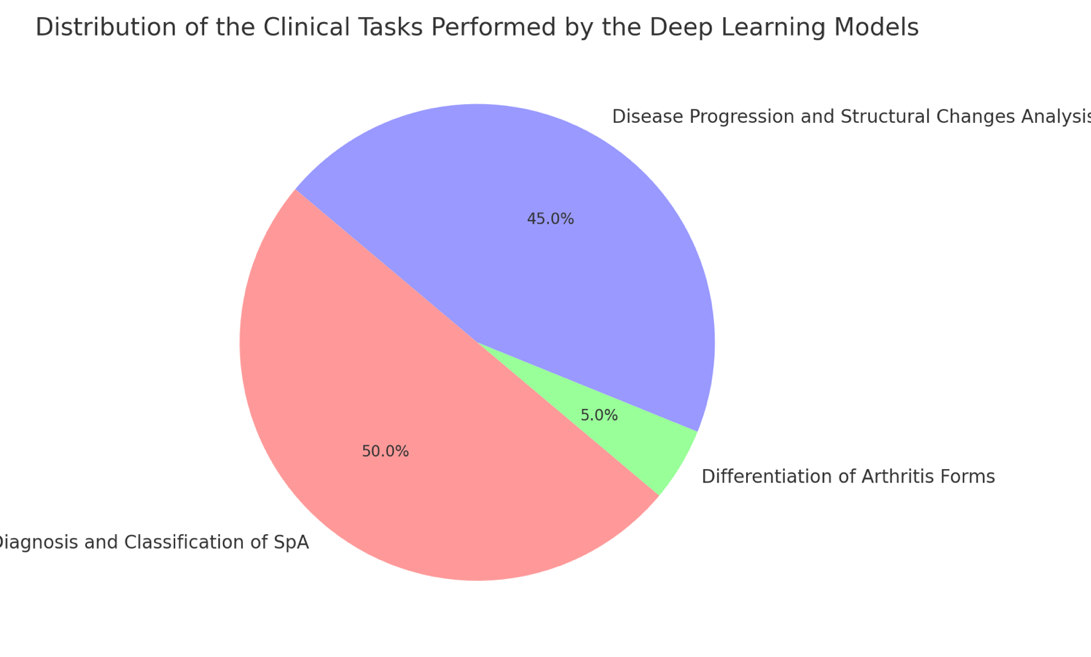
