## Supplementary material for "The Role of Deep Learning in Diagnostic Imaging of Spondyloarthropathies: A Systematic Review": Tables

**Table 1: A summary of the included studies.**

| Author | Year | Sample Size (Patients/Images) | Radiology method | AI Model | Clinical Task | Performance Metrics |
| --- | --- | --- | --- | --- | --- | --- |
| Lee (17) | 2023 | 296/4746 MRI slices | MRI | Faster R-CNN, VGG-19 | Detection of inflammatory sacroiliitis | Sensitivity: 0.725, Specificity: 0.936, AUC: 0.830 |
| Folle (18) | 2022 | 649 patients | MRI | ResNet neural networks | Differentiation of arthritis types | AUC: 0.67 - 0.75 |
| Bressem (38) | 2022 | 593 patients | MRI | 3D U-Net, ResNet-101 | Detecting axSpA changes in sacroiliac joint | Sensitivity: 88%, Specificity: 71% |
| Koo (37) | 2022 | 1280/10328 radiographs | Digital Radiography (X-ray) | Modified HRNet, ResNet 152 | Grading of vertebral bodies in AS | Sensitivity: 0.93652, Specificity: 0.97266 |
| Bordner (36) | 2023 | 256/362 MRI exams | MRI | Mask-RCNN | Predicting active sacroiliitis | MCC: 0.90, AUC: 0.98 |
| Roles (35) | 2023 | 279/243 patients | MRI | ResNet18-based CNN | Predicting Bone Marrow Edema | Cross-validation AUC: 94.5%, Balanced Accuracy: 80.5% |
| Gou (34) | 2021 | 100 subjects | MRI | LHR-Net, ResNet-50-based classification network | Segmentation and grading of AS lesions | DSC: 0.71 |
| Lee (33) | 2021 | 60/815 MRI images | MRI | ResNet18-based CNN | Detecting bone marrow edema | Accuracy: 93.55%, Recall: 92.87% |
| Zhang (9) | 2024 | 485 patients | MRI | ResNet50, ResNet101, DenseNet121 | Diagnosing sacroiliitis in axSpA | AUC: 0.839, Accuracy: 0.804 |
| Zhang (15) | 2023 | 435 CT exams | CT | nnU-Net, 3D CNN | Segmenting and grading sacroiliitis in AS | Dice Coefficients: 0.915, 0.889 |
| Lee (31) | 2023 | 492 patients | X-ray | DenseNet121 CNN | Diagnosing sacroiliitis | Sensitivities and Specificities peaking at 100% |
| Berghe (32) | 2023 | 145 patients | CT | U-Net, CNNs for erosion and ankylosis detection | Detecting structural lesions of sacroiliitis | Dice Coefficient for segmentation: 0.75 |
| Tenório (29) | 2022 | 46 MRI exams | MRI | ANN for radiomic model | MRI-based radiomic feature identification | Sensitivity: 100%, Specificity: 92% |
| Shenkman (30) | 2019 | 242 CT scans | CT | U-Net classifier, random forest | Diagnosing sacroiliitis | Sensitivity: 95%, AUC: 0.97 |
| Bressem (8) | 2021 | 1553/458 radiographs | Conventional Radiography | ResNet-50 CNN | Detection of radiographic sacroiliitis | AUC: 0.97, Sensitivity: 88%, Specificity: 95% |
| Li (7) | 2023 | 6436 PXRs | Pelvic Radiographs (X-ray) | Ensemble deep learning models | Diagnosing and predicting ankylosing spondylitis | Precision: 0.91, Recall: 0.90, AUC: 0.96 |
| Tas (20) | 2023 | 527 patients | MRI | DenseNet201 with GAP layer and kNN classifier | Diagnosing ankylosing spondylitis | F1-scores: 99.80% - 99.45% |
| Ureten (19) | 2023 | 585 radiographs | Pelvic Radiographs (X-ray) | CNNs with transfer learning | Diagnosing sacroiliitis | Accuracy: 89.9%, Sensitivity: 90.9%, Specificity: 88.9% |
| Rzecki (28) | 2021 | 30 MRI exams | MRI | U-Net-like architecture, VGG-like networks | Detecting bone marrow oedema lesions | Sensitivity: 0.88, Specificity: 0.91 |
| Fernandez (6) | 2022 | 267/534 radiographs | Conventional Radiographs (X-ray) | CNN-XGBoost model | Classification of sacroiliitis grade | Accuracy: 57%, Sensitivity for all classes except Class 1: over 60% |
| Lin (27) | 2024 | 330 patients | MRI | Attention U-Net algorithm | Detecting spinal inflammation in axSpA | Sensitivity: 0.80, Specificity: 0.88 |
| Faleiros (26) | 2020 | 56 MRI exams | MRI | SVM, MLP, Instance-Based Algorithm | Classifying active inflammatory sacroiliitis | Sensitivity: 100%, Specificity: 95.6% |

**Abbreviations:** MRI: Magnetic Resonance Imaging | CT: Computed Tomography | CNN: Convolutional Neural Network | ANN: Artificial Neural Network | SVM: Support Vector Machine | MLP: Multilayer Perceptron | AS: Ankylosing Spondylitis | axSpA: Axial Spondyloarthritis | PXRs: Pelvic Radiographs | GAP: Global Average Pooling | kNN: k-Nearest Neighbors | MDARG: Multi-Dimensional Features Automatic Radiomics Grading Algorithm | AUROC: Area Under the Receiver Operating Characteristic Curve | AUC: Area Under the Curve | MCC: Matthews Correlation Coefficient | DSC: Dice Similarity Coefficient.

**Table 2: Key Findings, Implications and Challenges of the included studies.**

| Author | Main Findings | Implications | Challenges |
| --- | --- | --- | --- |
| Lee (17) | Improved detection of inflammatory sacroiliitis using a two-stage AI model | Potential in enhancing MRI accuracy and reducing inter-observer variability in clinical settings | Reliance on expert consensus for ground truth, lack of real-world testing |
| Folle (18) | AI effectively differentiates between PsA, seropositive RA, and seronegative RA using MRI of the hand | Potential in differential diagnosis of arthritis forms using MRI scans | The need for larger training datasets |
| Bressem (38) | High sensitivity and specificity for detecting axSpA changes in sacroiliac joint with 3D U-Net and ResNet-101 | Potential in aiding axial spondyloarthritis diagnosis, especially in distinguishing between inflammatory and structural changes | Potential biases due to exclusive use of semicoronal images |
| Koo (37) | High performance in grading vertebral bodies, aiding in the detection of mSASSS in AS patients | Suggests deep learning's role in enhancing radiographic assessment of ankylosing spondylitis | Exclusion of cases with severe malformations, risk of overfitting, lack of clinical verification |
| Bordner (36) | High accuracy in detecting bone marrow edema and predicting active sacroiliitis using Mask-RCNN | Indicates potential for diagnosing sacroiliitis, particularly in axial spondyloarthritis context | Exclusion of MRI exams due to inadequate protocol |
| Roles (35) | Effective prediction of Bone Marrow Edema using ResNet18-based CNN, with high AUC in both validation and test datasets | Underlines the potential of automated ML pipeline in standardizing the evaluation of BME in MRI for spondyloarthritis | Challenges of class imbalance, difficulty in predicting underrepresented inflammatory lesions |
| Gou (34) | LHR-Net demonstrated superior lesion segmentation and grading performance in AS diagnosis | Highlights AI's significant potential in enhancing radiographic diagnosis of SpA | Data variability, generalizability across different medical imaging scenarios |
| Lee (33) | High accuracy in detecting bone marrow edema from MRI images using ResNet-based CNN | Demonstrates AI's effectiveness in aiding axial spondyloarthritis diagnosis, leading to improved detection and management | Lack of external validation, potential for selection bias |
| Zhang (9) | Excellent diagnostic performance in diagnosing axSpA-related sacroiliitis using CNNs and ensemble models | Shows robust potential of DLR combined with clinical factors in diagnosing sacroiliitis, advancing AI's role in radiographic diagnosis | Limited to oblique coronal MRI images |
| Zhang (15) | High segmentation accuracy and reliable grading of sacroiliitis on CT images using nnU-Net and 3D CNN | Demonstrates potential of deep learning for automated segmentation and grading of sacroiliitis in AS | No gold standard for sacroiliitis diagnosis, small dataset with potential bias |
| Lee (31) | High accuracy in detecting and grading sacroiliitis using DenseNet121 CNN on X-ray images | Diagnosing SpA, impacting treatment strategies and patient outcomes | Variability in image quality, patient positioning |
| Berghe (32) | Effective detection of structural lesions of sacroiliitis on pelvic CT scans using U-Net and CNNs | Suggests AI's significant role in diagnosing sacroiliitis, leading to earlier detection and treatment, and integrated diagnosis with clinical data | Limited number of pelvic CTs, focus on tertiary university hospitals |
| Tenório (29) | MRI-based radiomics correlated with Spondyloarthritis, with diagnostic performance comparable to radiologists | Indicates potential of MRI-based radiomics in supporting clinical assessment and differentiating axial and peripheral SpA | Small sample size and manual segmentation process |
| Shenkman (30) | AI model accurately detects and grades sacroiliitis, assisting radiologists in more efficient diagnosis. | Enhances diagnostic processes in radiology, indicating AI's role in improving efficiency and accuracy. | Potential for selection bias |
| Bressem (8) | Near expert-level detection of radiographic sacroiliitis with high agreement with human readers. | Supports potential use of AI for accurate sacroiliitis detection, beneficial in various clinical settings. | Patients already diagnosed with axSpA, unknown performance in undiagnosed patients |
| Li (7) | Superior diagnostic capabilities of ensemble DL models in diagnosing ankylosing spondylitis, effective even with smartphone-captured images. | Suggests AI can enhance the diagnostic process for ankylosing spondylitis, especially in areas with limited specialized care. | Difficulty in amassing large-scale PXR dataset |
| Tas (20) | High accuracy, recall, precision, and F1-scores in diagnosing ankylosing spondylitis using DenseNet201 with GAP layer and kNN. | Implies AI models like ASNET could improve ankylosing spondylitis diagnosis accuracy and healthcare resource efficiency. | Lack of external validation |
| Ureten (19) | Promising results from CNN models in diagnosing sacroiliitis from pelvic radiographs, with VGG-16 showing slight superiority. | Demonstrates potential of deep learning methods in aiding sacroiliitis diagnosis, possibly reducing reliance on advanced imaging like MRI. | Manual cropping of images, limited dataset size, lack of classification according to modified New York criteria |
| Rzecki (28) | High precision in automated detection and volume assessment of inflammatory lesions in axial spondyloarthritis. | Suggests significant potential for AI in enhancing diagnosis accuracy and efficiency, especially in early detection of bone marrow edema lesions. | Challenges in training deep learning models to distinguish closely located bones, reliance on expert knowledge for manual segmentation |
| Fernandez (6) | AI model effectively detects the grade of sacroiliitis on conventional radiographs with over 60% accuracy. | Highlights AI's aid in detecting radiographic sacroiliitis, assisting non-expert clinicians in diagnosis and reducing delays. | Complex dataset, small number of images per class, complete misclassification of Class 1 |
| Lin (27) | AI model shows comparable sensitivity and specificity to radiologist in identifying spinal inflammation in axSpA. | Indicates potential of AI in improving MRI interpretation for axial spondyloarthritis, aiding clinical management and diagnosis. | Potential bias in establishing ground-truth masks, small sample size, focus on identification over precise outlining |
| Faleiros (26) | Machine learning methods, particularly MLP, accurately classify active inflammatory sacroiliitis in MRI images. | Suggests AI can significantly aid in diagnosing active inflammatory sacroiliitis, improving early detection and treatment. | Small sample, segmentation by one radiologist, manual selection of images |

**Abbreviations:** AI: Artificial Intelligence | axSpA: Axial Spondyloarthritis | AS: Ankylosing Spondylitis | MRI: Magnetic Resonance Imaging | CT: Computed Tomography | CNN: Convolutional Neural Network | MLP: Multilayer Perceptron | PsA: Psoriatic Arthritis | RA: Rheumatoid Arthritis | mSASSS: Modified Stoke Ankylosing Spondylitis Spinal Score | BME: Bone Marrow Edema | LHR-Net: Lightweight Hybrid Multi-scale Convolutional Neural Network | DLR: Deep Learning Radiomics | nnU-Net: No-new-UNet | MDARG: Multi-Dimensional Features Automatic Radiomics Grading Algorithm | GAP: Global Average Pooling | kNN: k-Nearest Neighbors | PXRs: Pelvic Radiographs.
